# The Medicine Quality Monitoring Globe: A public health intelligence system for collecting and curating reports of substandard and falsified medical products from publicly available Internet news media

**DOI:** 10.64898/2025.12.19.25342665

**Authors:** Clark C. Freifeld, Kerlijn Van Assche, Alberto Olliaro, Martial Battain, Inthapavanh Kitignavong, Vayouly Vidhamaly, Phonepasith Boupha, Kem Boutsamay, Ngan Thi Do, Konnie Bellingham, Viengsavanh Pimxsayvong, Thitthiphone Olinh, Ana Rosado Olmo, Jingying Xu, Shan Jiang, Andrea Soult, Philippe J. Guerin, Paul N Newton, Céline Caillet

**Author notes:** These authors contributed equally to the work (Paul N Newton, Céline Caillet).

## Abstract

**Background:** The circulation of substandard and falsified (SF) medical products, including medicines, vaccines, and medical devices, continues to impact health of populations worldwide. Meanwhile, surveillance of SF products and data sharing are limited in most of the world, and incidents are rarely published in accessible databases. We describe here the Medicine Quality Monitoring Globe system (MQM Globe) for capturing, curating, characterising, and disseminating online media reports relating to SF products. The system consists of the curation environment and the Web application.

**Methods:** Online reports are acquired from search engines, direct feeds, and targeted extraction, in five languages. They then pass a series of automated content characterisation steps to filter and label them by relevance, location, and product. Reports are then reviewed by a trained curator staff, before being made available through a visualisation Web application.

**Findings:** Over the study period from July 2018 through December 2024, we identified 11,000 relevant distinct SF incidents, across a broad range of geography, products, and categories. The automated relevance classifier exhibited precision of 37%, recall 90%, and F1-score 52%, for English-language content.

**Interpretation:** The information captured by the MQM Globe is publicly available and can help inform public health authorities as situations emerge. It is the only publicly available repository of open source intelligence specifically on SF medical products. It can be used as an informational tool to trigger intelligence-led activities across law enforcement, customs and Medicine Regulatory Authorities.

**Funding:** This research was funded in whole, or in part, by the Wellcome Trust [202935/Z/16/Z and 106698/Z/14/Z]. The authors have applied a CC BY public copyright licence to any Author Accepted Manuscript version arising from this submission. The Gates Foundation supported the development of the Regulatory and Alerts webpage and the adaptation of the Globe to respond to SF COVID-19 medical products.

**Research In Context:** *Evidence before this study:* The World Health Organization estimated that approximately 10.5% of medicines in low- and middle-income countries are substandard or falsified (SF). However, there is limited evidence on the extent of the problem, due to its nature, the weak regulation capacity in many poor-resourced countries, the complexity of global supply chains, and lack of awareness on the issue. To date there has been little analysis of publicly available online media reporting on events related to SF medical products.

*Added value of this study:* This article describes the development and assessment of a publicly available online platform, the Medicine Quality Monitoring Globe, for monitoring news media and regulatory agency information related to SF medical products events. This active and timely system curates and organises reports in five languages.

*Implications of all the available evidence:* This article introduces a tool that offers a new source of data for better understanding the global challenges related to SF medicinal products, and serves as an early warning system for incidents that may require investigation and action from regulatory authorities, health professionals, researchers and the public.

## Introduction

The widespread circulation of substandard and falsified (SF) medical products, including medicines, vaccines, and medical devices, continues to cause substantial morbidity and mortality worldwide. The World Health Organisation (WHO) estimated that 10.5% of medicines in lower- and middle-income countries are SF, and wealthier countries are by no means immune (WHO, 2017). The WHO and other international organisations, along with national medicines regulatory agencies, have made strides in establishing common terminology, understanding the scope of the problem, and initiating collaborations to address it (WHO, 2017; Newton, Bond et al, Lancet Global Health 2019). Falsified medical products are defined by the WHO as those “that deliberately/fraudulently misrepresent their identity, composition or source,” in contrast to substandard or “out of specification” medical products that “are authorised medical products that fail to meet either their quality standards or their specifications, or both” (WHO, 2017). Both research and innovative systems on the epidemiology of SF medical products, their detection, and their impact are neglected aspects of the growing field of public health security.

The extent of the problem is difficult to measure as most often the quality of a given medical product cannot be determined without specialised equipment and expertise in the relatively few reference laboratories globally. Furthermore, surveillance of SF products and data sharing are limited in most of the world, and incidents are rarely published in accessible databases or scientific journals. However, the broad adoption of the Internet and electronic media over recent decades presents an opportunity to monitor reports relating to SF medical products using software tools to provide early warning of potential problems.

We describe here the Medicine Quality Monitoring Globe system (MQM Globe) for capturing, curating, characterising, and disseminating online news media reports relating to SF medical products. The system consists of the curation environment (back-end and front-end) and the MQM Globe web app (back-end and front-end). The aim of this open-source intelligence (OSINT) system is to bring diverse multilingual news reports together, collate and curate them and facilitate their access, to provide timely warning of potential problems with SF products for national and international organisations, and enhance policy discussion and public dissemination of these data.

To date, global sharing of data on SF medical products has been limited (Cockburn et al., 2005)(Cockburn et al. 2005, Newton et al. 2014, Access to Medicines Index 2024) due to commercial and political sensitivities that sometimes inappropriately override public health concerns. If local, regional, and global data on SF incidents are more accessible to the public, academics, journalists, and regulatory bodies, pressure may be brought to improve appropriate data sharing and timely, appropriate interventions, enhancing public health security. In our understanding, the MQM Globe is the only publicly available repository of open source intelligence specifically on SF medical products.

The MQM Globe shares information in two ways: the ‘lay press articles’ displaying online news reports, and ‘regulatory & alert webpages’ that lists the medicines regulatory authorities (MRA) webpages of recalls/warnings/alerts on SF medical products. We describe in this article how the system was developed, how it works, and ideas for how it could be improved and sustained.

## Methods

The MQM Globe has been publicly and freely available at https://www.iddo.org/mqmglobe/ since the 30^th^ of March 2020. It was inspired by the HealthMap (Brownstein & Freifeld, 2007) (Brownstein et al. 2007, 2008) and RadioGarden systems (Radio Garden, 2025).

The curation environment consists of a back-end with a data acquisition pipeline and various facets of automated labelling of the data, and a front-end with a curation and labelling interface for use by trained analysts. Once the data are characterised, they are made publicly available through the MQM Globe web app front-end system, a search and visualisation tool. We present here the details of the implementation of each of these components (**Figure 1**).

**Figure 1.**
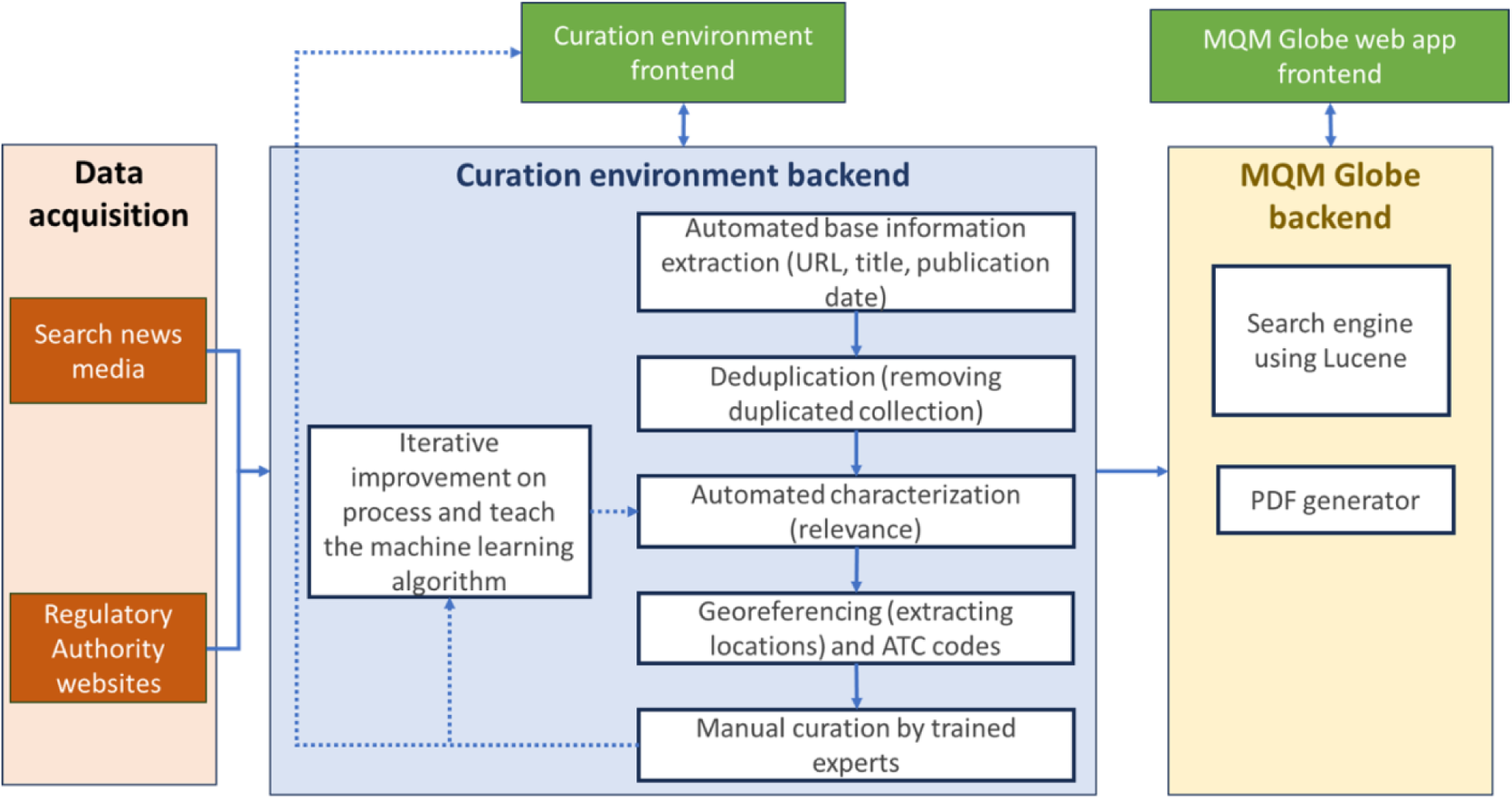
Workflow of the Medicine Quality Monitoring Globe system (MQM Globe): from data sources to the web interface.

Engagement with key stakeholders, such as the WHO, Europol, Interpol, national MRA, and other researchers in this field was conducted during the planning and construction of the system, such as at the Medicine Quality & Public Health Conference in September 2018 (Centre for Tropical Medicine and Global Health, 2018). The MQM Globe release (Centre for Tropical Medicine and Global Health, 2020) was disseminated widely through fora for global internet community of practice on access to essential medicines e-drug (English) and e-med (French) (Newton, 2020) and international collaborators through the Medicine Quality Research Group network.

### The curation environment – collecting and curating data from online news reports

#### a. Data acquisition

The MQM Globe system acquires online news media reports from search engines, direct RSS (Really Simple Syndication) feeds, and targeted “web scraping” (parsing and extraction from HTML).

Most of the reports identified via search engines come from Google News, using specific key terms adapted for each language (**Supplementary material 1**). Because Google News restricts the number of key terms that can be used per query, multiple search queries are used. Each query combines one term related to a medical product and all terms and synonyms related to quality (e.g. falsified, substandard, counterfeit, fake). The system also uses exclusion terms selected by language to exclude commonly occurring false positives identified during the testing of the first iteration of the system (e.g. “fake news”) (**Supplementary material 1**). The searches are performed through an automated system, multiple times per day. For each search query, Google News returns results in XML (eXtensible Markup Language) format which are then parsed and processed by the system.

The development was conducted initially for English language articles, and four other languages were added later (French, Spanish, Vietnamese, and Mandarin Chinese). For each language, key terms were provided by translation of the English terms and synonyms, where applicable, by native language speakers. They have been regularly revised since.

The system also uses the Baidu search engine (Baidu, 2025) for searching news media in Mandarin language, using three queries. As Baidu does not provide a structured machine-readable result, the system therefore parses the HTML results and converts them to structured form.

In addition to the news processing pipeline of the search engines, the system captures reports from RSS feeds and sources that require “web scraping” (**Supplementary material 3)**. For each collected report, the acquisition process produces a standardized set of data fields, such as the report’s publication date, title, and URL (Uniform Resource Locator) from the diverse array of sources, and extracts plain text from the original report HTML using the *textract* library (Bashford D, 2018).

The system was tested in 2016 and 2017 in English, and between 2018 and 2019 in other languages, to refine search terms and adapt the interface and the data extracted. The MQM Globe Web front end displays data from July 2018 for English, February 2019 for French, June 2019 for Mandarin Chinese, August 2019 for Spanish, and September 2019 for Vietnamese.

#### b. Deduplication

Since July 2021, the system has been performing an automated deduplication process based on report title to avoid displaying the same report published in different sources multiple times. The algorithm considers two reports duplicate if their headlines are 90% the same text. For all reports that are flagged as relevant, the analysts perform an additional manual screening for duplicates. There is currently no system to identify cross-language duplicates.

#### c. Automated characterisation

Once the reports are loaded, the system applies automated characterisation of (i) the relevance of the report, (ii) geographical information (georeferencing) for English language, (iii) products mentioned using the ‘Anatomical, Therapeutic and Chemical’ (ATC) classification for English, French and Spanish languages, and (iv) duplication with respect to previously collected articles.

##### i. Relevance classifier

The system employs a machine learning model to mark news reports for relevance since 2018 for the English language, 2019 for the French language, 2024 for the Spanish and Vietnamese, and 2025 for Mandarin Chinese. This automated ‘relevance classifier’ is a binary classifier, labelling each report as relevant (hereafter ‘positive’), or not relevant (hereafter ‘negative’). The system is calibrated to have approximately 90% recall to limit false negatives (see section **Performance of Automated Characterisation** below). The downside of missing a true positive is deemed greater than the downside of having to re-label a false positive. The model is based on a Fisher-Robinson classifier (Robinson G, 2003), implemented in Python and using MongoDB to store the model data.

##### ii. Georeferencing

The system uses the CLIFF-CLAVIN software package (D’Ignazio et al. 2014) to identify the main locations linked to the incident described in each report, at the highest relevant geographical resolution, generally a city or town. As CLIFF is limited to English-language content, the automated characterisation is only applied to English-language reports.

##### Iii. ATC codes

To identify product mentions, in English, French, and Spanish, the curation back-end uses a dictionary matching algorithm based on a Trie data structure (de la Briandais, 1959), built from the WHO - ATC standard (WHO, 2024). The ATC system classifies active pharmaceutical ingredient (API) and combinations of API into different groups according to the organ or system on which they act and their therapeutic, pharmacological, and chemical properties. For the automated characterisation, all different levels of the ATC classification are included.

#### d. Analyst Labelling

The reports collected by the back-end environment are accessed by analysts through a web-based front-end curation tool with restricted access. The analysts are responsible for the curation of the data for the articles automatically labelled as relevant by the relevance classifier. This includes the validation of the relevance, where necessary the correction of automated characterisation data, and the extraction of the information that has no initial automated step, namely: the quality type (e.g., substandard, falsified), medical product source (i.e., where the medical products were identified/seized), categories (e.g., antibiotics, veterinary medicines), incident date, and an extract of essential information (e.g. name and batch numbers of medicines, description of quality issues, if available).

Data analysts for each language were trained to curate the data according to standard operating procedures (available on request to the authors). Reports on SF medical products without specific incidents are only validated for their relevance and labelled as ‘General discussion’ without further processing (see MQM <u>Globe site</u> for more information).

### Visualisation on the Medicine Quality Monitoring Globe web application

#### a. Overview

The MQM Globe Web app consists of two main components: a public-facing front-end interface written in Angular (version 6) combined with Cesium, a JavaScript library for 3D geospatial applications, and a back-end interface developed in Java and Lucene that provides, through a set of Web services, data to the front-end.

The MQM Globe is divided as follows: the left-side panel containing the ‘Lay press articles’ tab (**Fig. 2 section 1**), opened by default, and the ‘Regulatory & alert webpages’ tab (**Fig. 2 section 2**) at the top; the large central panel displaying a globe (**Fig. 2 section 3**) with incidents geographically placed; the right-hand panel “Medicine Quality Monitoring Globe Index” (**Fig. 2 section 4**) menu links to documentation including user guide, tutorial video, methodology, acknowledgments, and disclaimers and caveats. On the top right of the central panel, users can change the view, e.g. transforming the globe from the default 3D view into a 2D map (**Fig. 2. section 5**).

**Figure 2.**
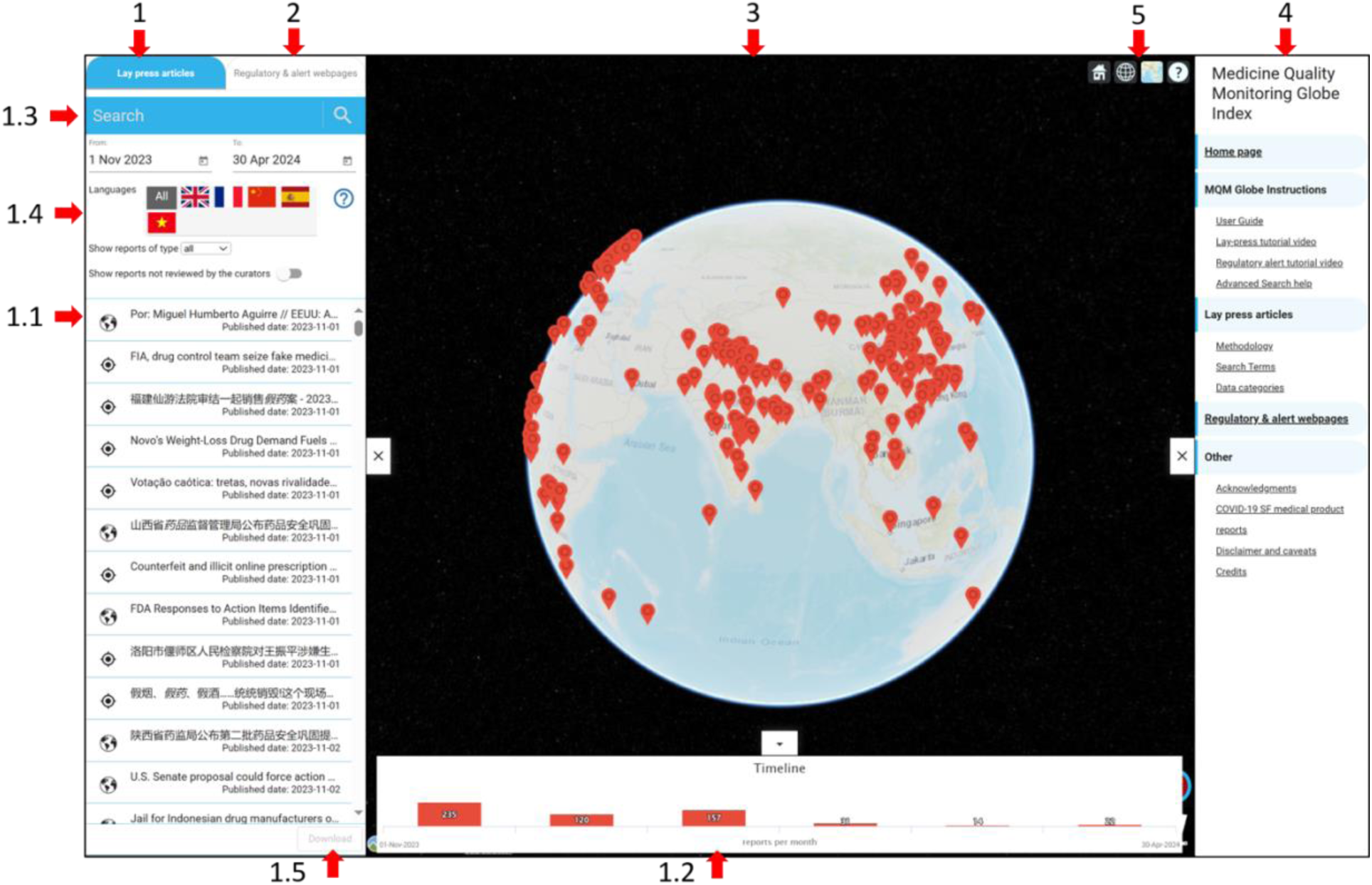
The ‘Medicine Quality Monitoring Globe’ (MQM Globe) front-end. The four main sections are indicated 1) ‘Lay press articles’ tab, 2) ‘Regulatory & alert webpages’ tab, 3) Visual of the geographically placed incidents, 4) Index and hyperlinks to documentation. Panel 1.1 to 1.5 are features linked to the ‘Lay press articles’ tab. Available at: https://iddo.org/mqmglobe

#### b. The ‘Lay press articles’ tab

##### i. Overview

On opening the ‘Lay press articles’ tab, reports for the previous three months are displayed on a 3D map (**Fig. 2 panel 3**) as well as in list format inside the left-side panel (**Fig. 2 section 1.1**). Only reports that describe incidents involving suspected SF medical products such as recalls, seizures, alerts, or case reports of an unexpected effect for a patient, with a geographical location, are represented by pins. Clicking on a pin opens a popup showing further details, including links to the source reports (**Fig. 3**), and information curated by the analysts: the ATC codes, quality, source, and medicines category. One pin can represent one or more reports.

**Figure 3.**
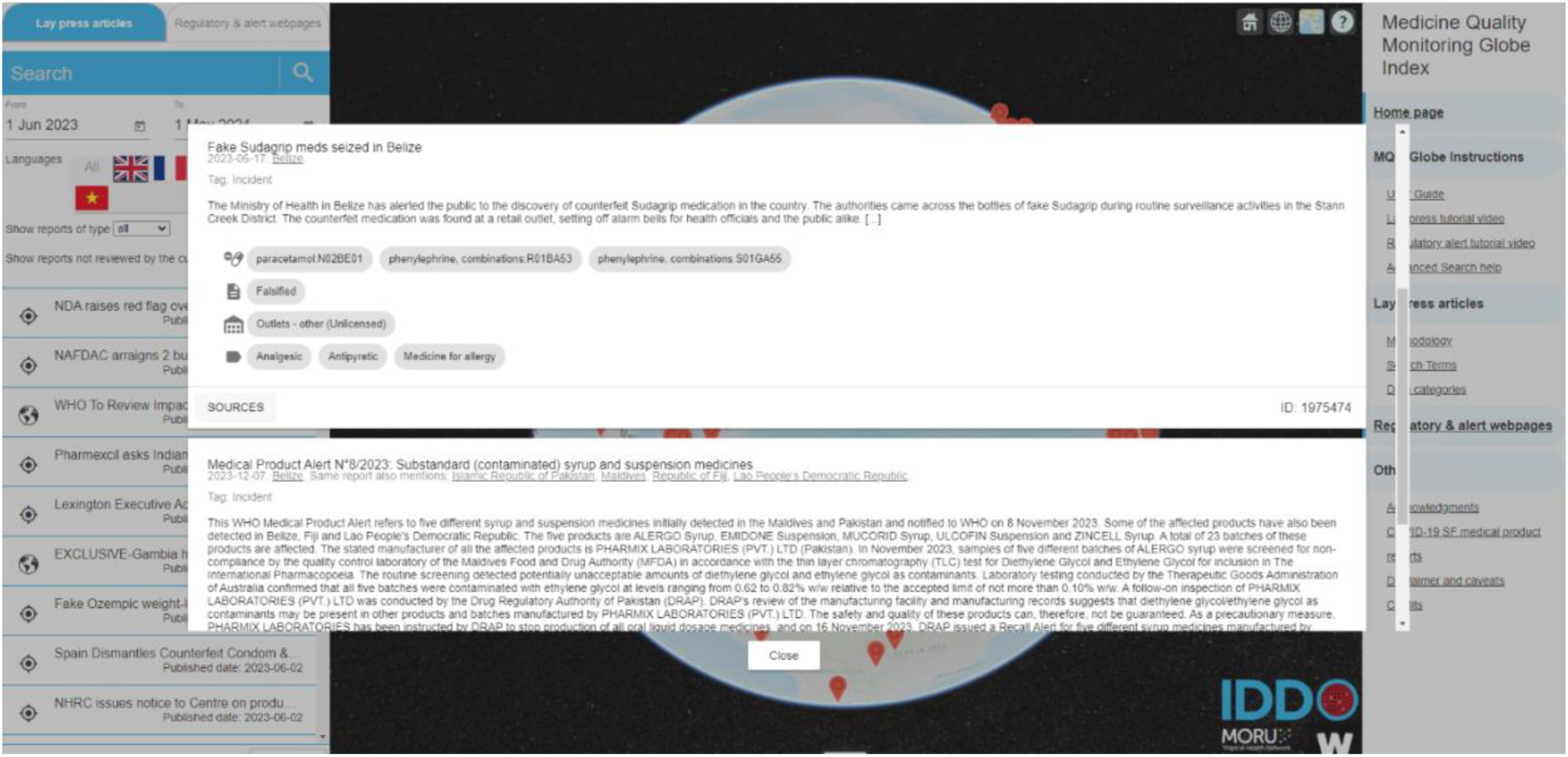
An example of the information displayed when clicking on an incident pin on the Medicine Quality Monitoring Globe front-end. Available at: https://iddo.org/mqmglobe.

Reports describing incidents are also represented in the search results column of the left-hand panel with a target icon. In that list, general discussions on medical product quality issues without description of incidents are also recorded and marked by a globe icon. More details on the type of reports and incidents included in the system are available in the standard operating procedures (available on request) and on <u>the Globe site</u>.

The bottom “Timeline” panel (**Fig. 2 panel 1.2**) provides a visual representation of the temporal distribution of the reports as a bar chart (per year/month/week depending on date range). Clicking on one of the bars opens a scrollable list of all reports for that period.

##### ii. Searching the MQM Globe

To conduct a search, a user will typically enter terms into the search dialog box, such as a medical product or location of interest (**Fig. 2 panel 1.3**). In addition, the user can filter the results by date range, language of the original press reports (**Fig. 2 panel 1.4**), type of report (incident reports vs general discussion), and whether the data has already been or is yet to be reviewed by the curators. Clicking the magnifying glass will trigger the search by sending a request to the MQM Globe back-end. On receipt of the request, the service performs a free text search of database content using the Lucene index and filters according to the supplied additional parameters. The frontend will then display the updated results as pins on the Globe and in the list below the filters.

Users can download the data as a Portable Document Format (PDF) report in tranches of 1-100 reports (**Fig.2 panel 1.5)**. For all languages, to maximise the effectiveness of the search, ontologies are employed during the indexing of the data. For example, if a user types “Africa,” search will return reports that include, for example, Ethiopia or any other African country name in the location fields (more examples in **Supplementary Material 4**). The dictionary scheme holds several tables containing this taxonomic data, including synonyms, country names, and continents, in all 5 languages.

##### iii. COVID Shortcut Searches

To facilitate access to specific types of SF COVID-19 medical products (e.g. COVID diagnostics, Personal Protective Equipment), ‘Shortcut queries’ (**Supplementary material 2**) were added during the COVID-19 pandemic and can be accessed by clicking on the search box.

#### c. The ‘Regulatory and alerts web pages’ tab

The ‘Regulatory & alert webpages’ tab links to national regulatory authority webpages of recalls/alerts/warnings/seizures on SF medical products, and webpages of national or international organisations with a mandate to report about SF products, such as the WHO (**Fig. 4**). Note that not all MRAs publish data concerning alerts and recalls, and there is currently no standard amongst those that do publish on the level of information included in those alerts or recalls.

**Figure 4.**
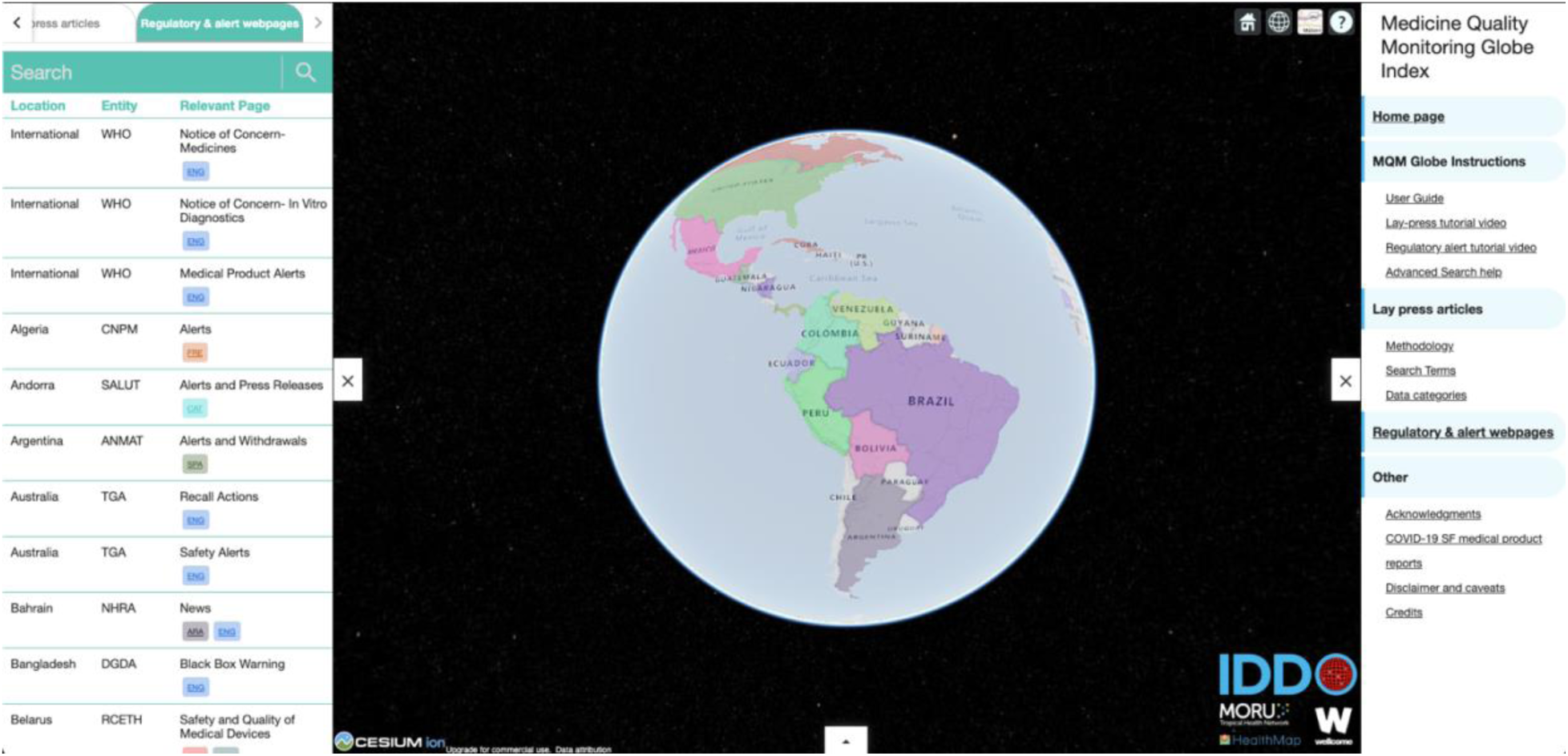
Screenshot of the ‘Regulatory & alert webpages’ tab-Medicine Quality Monitoring Globe front-end. Available at: https://iddo.org/mqmglobe. The tab contains a 3D geographical visual located in the centre, and a left-hand side panel showing the names of the entities who own the webpages, their country, and a brief description of the type of information in the webpage. Countries where the relevant webpages have been identified appear as coloured polygons. Clicking on a polygon triggers the listing of the relevant country’s webpages in the left-hand side panel.

## Results

### 1. Main characteristics of reports collected by the system

Between the system’s official inception (July 2018) and December 2024, a total of 2.3 million reports were captured (**Table 3**). Of these, 48,000 were determined to be relevant with 20,000 of those being distinct.

**Table 3:**
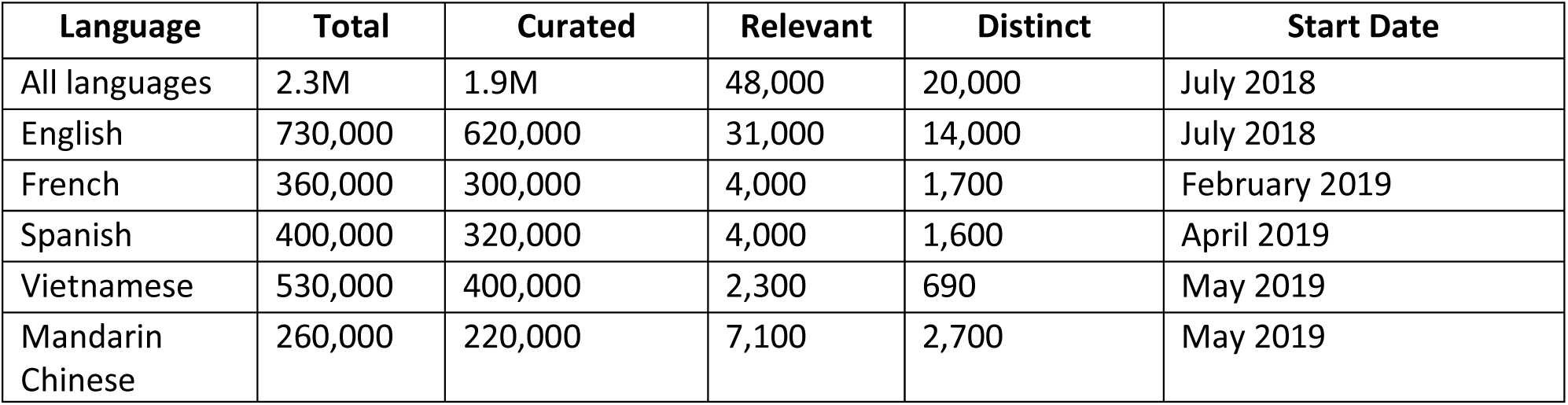
Number of reports collected by the system between July 2018 and December 2024, by source language.

Among the 20,000 distinct reports identified as relevant, 11,000 were labelled as ‘Incidents’ (i.e. shown on the Globe web app front-end), and 9,000 as ‘General discussions.’ The incidents were related to products that were mainly falsified (47.5%), substandard (21.0%), and diverted/unregistered (**Table 4**). The term ‘Substandard or Falsified’ was used when there was no indication, or uncertainty, regarding whether the products were substandard or falsified.

**Table 4:** Specific quality issues mentioned in incident reports and relative importance of each quality issue. Non-duplicate incident reports for all languages from July 2018 to December 2024.

| Quality issue | Number of reports | Percentage of quality issues |
| --- | --- | --- |
| Falsified | 5,249 | 47% |
| Substandard | 2,508 | 22% |
| Diverted/Unregistered | 1,571 | 14% |
| Substandard or Falsified* | 1,403 | 13% |
| Degraded | 191 | 2% |
| Unknown/Other | 306 | 3% |
| In total, 10,544 distinct reports describing at least one specific quality issue were identified. Hence, the sum of reports in the table may exceed the total number of distinct reports because certain categories can appear multiple times for a single report. |  |  |
| *There was no indication, or uncertainty, regarding whether the products were substandard or falsified. |  |  |

The most represented ATC entities (**Table 5**) include medicines often diverted for recreational use, such as the opioids oxycodone, fentanyl, or tramadol. Vaccines were part of the most identified ATC entities in all languages supported by an ATC dictionary.

**Table 5.**
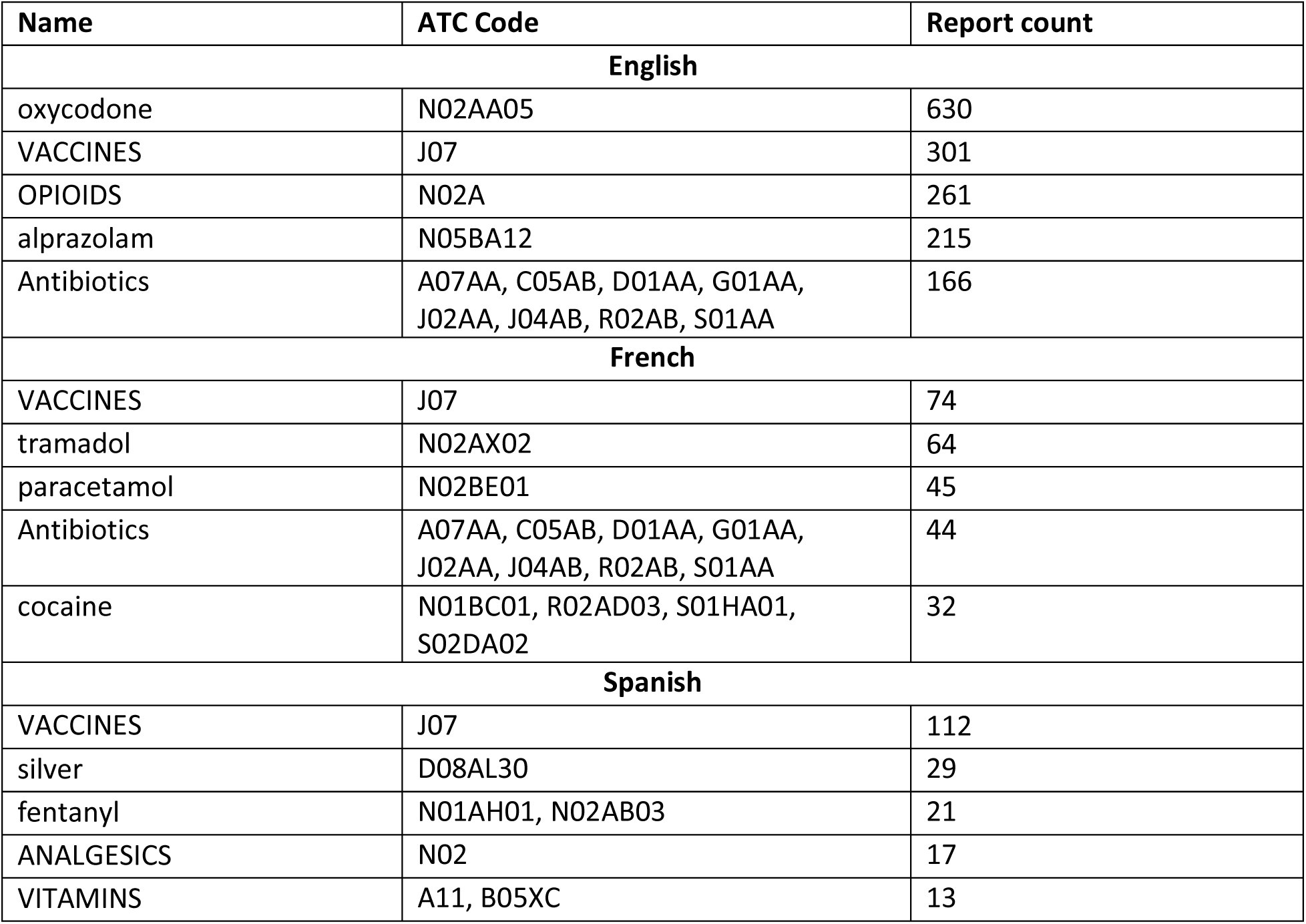
Top 5 most identified ATC entities in English, Spanish, and French. Non-duplicate incident reports from July 2018 to December 2024.

### 2. Performance of Automated Characterisation

During the initial years of the development, automated characterisation was confirmed or corrected by analysts. This allowed us to define positive and negative predictions and compute performance metrics, namely, precision, recall, and F1-score (harmonic mean of precision and recall).

#### Relevance classifier

We consider “positive” a report on SF medical products that describes an incident or is a general discussion. Over a sample of 15,000 English reports collected by the system in 2021, the automated classifier identified 552 true positives (correct identification of a report as relevant), 13,429 true negatives (correct identification of a report as not relevant), 955 false positives (wrong identification of a report as relevant), and 64 false negatives (wrong identification of a report as non-relevant), resulting in precision of 37%, recall 90%, and F1-score 52%. For French language, on a sample of 15,000 reports, 65 were true positives, 14,772 true negatives, 160 false positives, and 3 false negatives resulting in a precision of 29%, a recall of 96%, and an F1-score of 44%.

#### Deduplication

We consider “primary” a report characterized by the curators as not a duplicate of another report, and “duplicate” a report for which the curators identified earlier reports of the same event.

To save work, the analysts don’t review articles identified as duplicates after July 2021 when the deduplication system was implemented. Hence, we performed a retrospective evaluation on a separate sample of 1,165 reports from July to December 2018, where analysts manually labelled reports as primary or duplicate. Considering primary reports as “positives” and duplicates as “negatives,” the deduplication system identified 338 true positives, 568 false positives, 10 false negatives, and 249 true negatives, for a precision of 37%, a recall of 97%, and an F1-Score of 54%.

#### Georeferencing

On a sample of 2,624 relevant non-duplicate English news reports collected from August 2021 to December 2024, the CLIFF system identified 1,242 true positive locations (identification of the correct location), 2,869 false positives (identification of an incorrect location), and 3,202 false negatives (failure to identify a correct location), yielding 30% precision, 28% recall and an F1-Score of 29%.

#### ATC codes

Out of a sample of 2,693 relevant non-duplicate English-language reports collected from July 2021 to December 2024, the ATC-code classifier led to 1,646 true positives (identification of the correct API), 2,161 false positives (identification of an incorrect API), and 1,561 false negative records (non-identification of the correct API), yielding a precision of 43%, a recall of 51%, and an F1-Score of 47%. For French-language, 372 relevant and non-duplicate reports (same period), 213 true positives, 130 false positives, and 176 false negatives were observed, with a 62% precision, 55% recall, and 58% F1-score. For Spanish-language, 607 relevant and non-duplicate reports, 513 true positives, 210 false positives, and 224 false negatives were observed, with 71% precision, 70% recall, and 70% F1-score.

### 3. Usage of the Medicine Quality Monitoring Globe

From its launch in March 2020 until December 2024, 3,080 unique users performed searches on the MQM Globe (**Fig. 5**). The searches were performed in 126 countries, including 78 low-and-middle-income countries (LMICs). Most of the searches were performed by users in the USA (n=1069; 32%), the UK (n=525; 16%) and France (n=207; 6%) (Further information in **Supplementary material 5)**.

**Figure 5.**
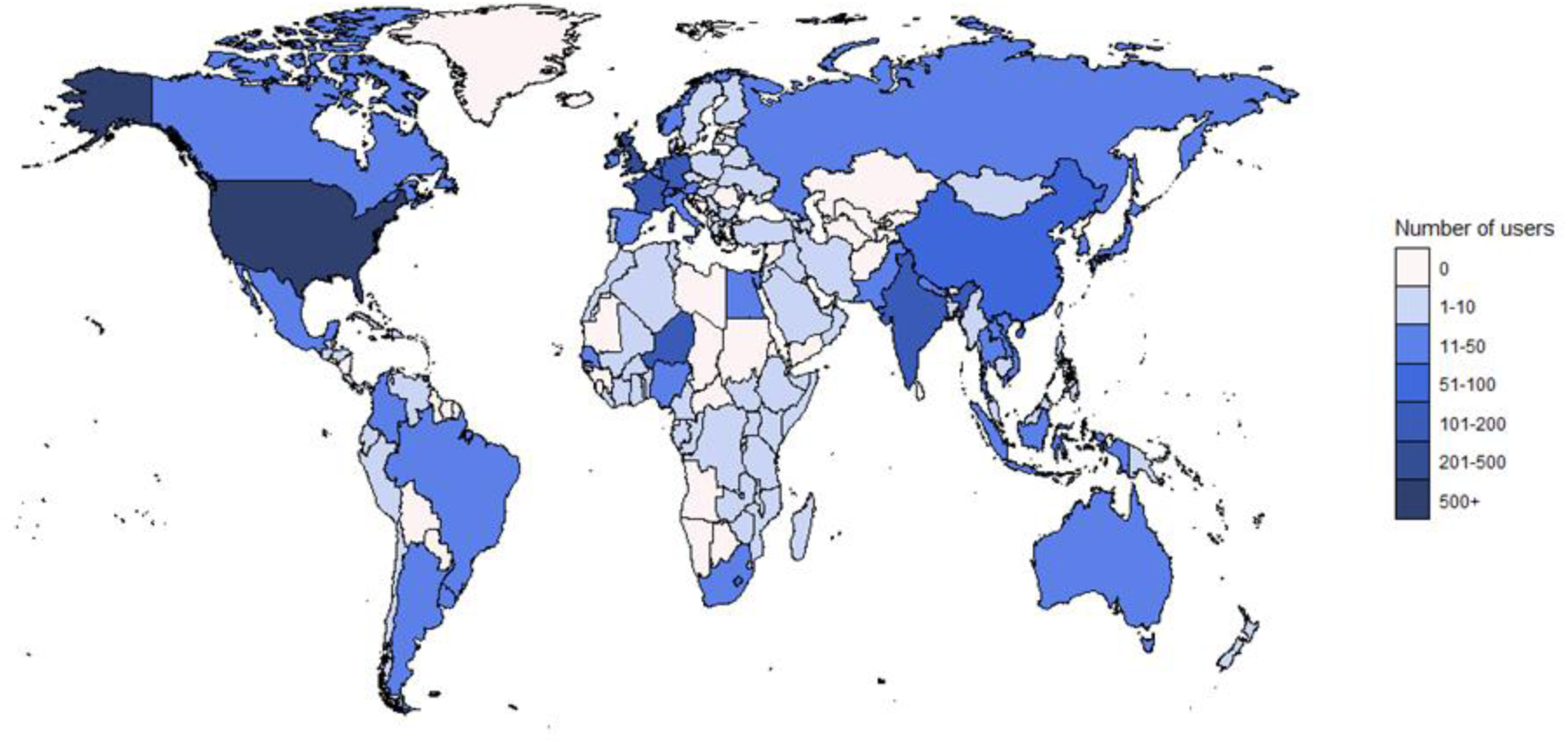
Distribution of unique users of the Medicine Quality Monitoring Globe.

## Discussion

We developed the MQM Globe, an interactive open access online tool, to map real-time media reports on poor-quality medical products worldwide. The MQM Globe provides customisable views of national and international online news for English, French, Spanish, Mandarin, and Vietnamese. The MQM Globe regulatory tab displays webpages of medicine regulatory agencies and international organisations.

The situation around children’s cough syrups containing diethylene glycol that emerged in India in 2020 demonstrates the Globe’s potential value as a tool for public health practitioners. The first related report appearing in the Globe database is an article from the Times of India from 23 February 2020, detailing how 17 children in Jammu were impacted, of whom 9 died (TNN, 2020). The WHO became aware of the issue only later, at the beginning of the summer 2025 (personal communication) when cases were identified in Africa. On 5 October 2022, the WHO issued the first global alert concerning paediatric cough syrups contaminated with diethylene glycol, suspected to have caused 66 deaths of children in The Gambia (WHO, 2022). This timeline shows the potential for the MQM Globe to act as a warning system to support further investigations.

As illustrated in this case, due to global supply chains, the risks of substandard and falsified medications are not constrained by geography. The information captured by the MQM Globe system is publicly available and can help inform public health authorities as situations emerge. For example, the MQM Globe can be of use to governments and procurement agencies in conducting due diligence prior to authorising imports or entering into procurement contracts, and should form part of their risk assessment process (a search on a company name can yield results that would lead to further research before awarding contracts). Law enforcement, customs, regulatory managers, and MRAs planning coordinated operations against SF medical products should consult the MQM Globe to influence intelligence-led activities. While the total volume of relevant, distinct reports identified through the Globe may be relatively small (11,000 over the period studied), reports usually refer to a batch of medicine. If falsified, the size of the batch is usually unknown, if substandard the batch should be known. In both cases, the sizes can range up to hundreds of thousands and sometimes millions of doses. The potential to adversely affect public health is large.

Meanwhile, beyond SF pharmaceuticals, there is increasing interest in the use of OSINT for public health to collate and disseminate early warnings of other issues that may need intervention, such as for food fraud (Bouzembrak & Marvin, 2016, Marvin et al. 2022), infectious diseases (Epidemic Intelligence from Open Sources, 2023) and the illegal wildlife trade (Patel et al. 2015).

The information included in the MQM Globe is extracted from diverse resources and responsibility for the accuracy of these data and their interpretation rests with the primary resource. A parallel platform, the ‘Medicine Quality Scientific Literature Surveyor’ (IDDO, 2025), collates and curates data from the scientific literature, that we expect will be more reliable and trustworthy than those data represented in the Globe, that will often not have scientific confirmation. Therefore, interpretation of the information in the Globe should be exercised with the greatest caution. However, these reports are early warnings of potential problems that need further investigation, if not already conducted. MQM Globe newspaper reports are analogous to pharmacovigilance patient adverse event reporting, such as the UK Yellow Card scheme (MHRA, 2025).

The MQM Globe data are geographically heterogeneous, and an important caveat is that no or few reports from an area does not imply that medical product quality there is good, but rather that there may be limited accessible reports. Similarly, many reports of poor-quality medical products in a country do not imply that medical product quality there is universally worse than elsewhere. Countries with many news reports should be lauded for facilitating such reporting. In addition, although the analysts endeavour to include published corrections of previously published news, we cannot guarantee that these are included. Moreover, there is often confusion in the lay literature between the terms falsified, substandard, spurious, fraudulent, degraded and counterfeit, and context is needed when interpreting these terms.

Overall performance of the automated relevance classifier for online news media reports within the curation environment is not as high as we would like; one of the challenges is the highly unbalanced nature of the data, with large numbers of irrelevant reports and few relevant reports; another challenge is the change of the technical infrastructure we rely on (The Google News platform constantly changes and evolves); yet another challenge is to be able to handle and classify emerging SF issues as they appear such as Covid19-related products or fentanyl. Though we have a large volume of labelled examples for use in our machine learning model, we have a relatively limited number of positive examples. The performance of the georeferencing system also has room for improvement, though the task is difficult given the vast number of possible named locations and the variability of the input (Ng et al. 2020). In future work, we may be able to boost precision through applying an improved post-processing step to the CLIFF output.

Large language model (Brown, 2020) tools are becoming more powerful and are promising for further development of the MQM Globe, and trials in our system are ongoing. LLMs may be able to assist with the deduplication of reports, their automated characterisation, georeferencing, and the attribution of the ATC codes. Online news media reports provide unstructured data that require considerable human effort to curate and extract the relevant information. Some of the curation fields that are currently filled manually, could be automated, enabling curators to handle larger quantities of information and reducing their workload.

Given the high number of user searches from high income countries (HIC), we are reinvigorating MQM Globe dissemination in LMIC, to facilitate uptake. As there are limited mechanisms for reaching out to medicines regulatory authorities globally, a publicly accessible reference database of points of contact for public or academic engagement would be a valuable asset for this purpose. To further improve the detection of SF product issues in different geographical areas, and reduce inclusion bias, more languages should be included in the system. For example, including Arabic and Russian would allow us to cover the six official languages of the UN.

The effectiveness of the system depends on detecting, retrieving, and compiling information from multiple sources. Currently we mainly rely on Google News as a news aggregator. Furthermore, while some of our other sources offer RSS feeds, information is often difficult to access and requires the development of “screen scraping” software to automate data acquisition. We suggest that regulatory authorities and non-governmental organisations who monitor and publicise events related to SF medicines offer machine-readable outputs to disseminate their findings more efficiently.

Data sharing on SF medical products has historically been woeful (Cockburn et al. 2005, Newton et al. 2014, Access to Medicines Index, 2024) and our aim is that the Globe, and the parallel Scientific Literature Surveyor, will assist in data sharing. For example, these tools could help countries to be alert to reports, albeit usually unverified, of SF medical products in neighbouring countries to inform their own risk-based post-market surveillance. Additionally, the Globe and Surveyor may help the public to further understand the problem and how it affects their communities to be able to argue, with evidence, for increased investment in regulatory oversight where needed.

The MQM Globe system needs further improvement through broader engagement with diverse users to understand their needs, as well as discussions on how to ensure its sustainability in the long term. Development of grading systems, if possible, to gauge the veracity of both source and information given in press reports, and to grade their public health risk, may help prioritise reports for urgent investigation.

A further sustainable iteration of this system with LLM facilitated curation and grading of veracity and risk, and the automatic transmission of reports to national and international regulatory authorities could help practical alert dissemination to inform rapid investigation and regulatory/enforcement interventions and encourage data sharing to protect public health security.

We hope that the current system assists with the prevention, detection, and response (WHO, 2017) to SF medical products, by facilitating access to diverse reports from around the globe at one interface. The intended audience is primarily national organisations such as medicines regulatory authorities and ministries of health and enforcement, international organisations such as WHO, UNODC, Europol, Interpol, and pharmaceutical manufacturers, distributors, and sellers. We welcome suggestions as to how to improve the system.

## Data Availability

All data produced in the present study are available upon reasonable request to the authors. Much of the data is available online at https://www.iddo.org/medicine-quality-monitoring-globe

## Acknowledgments

For the purpose of Open Access, the author has applied a CC BY public copyright licence to any Author Accepted Manuscript version arising from this submission.

We would like to acknowledge the members of the FORESFA Collaboration for diverse discussions and cooperation that benefitted this research.

This paper is dedicated to the memory of the late Dr. Andrew Payne, who expertly led the frontend development of the MQM Globe Web application and worked on the first draft of this paper.

## Supplementary materials

As of January 2025, there are a total of 161 queries for English language, 201 for French language, 182 for Spanish language, 170 for Vietnamese language, and 297 for Mandarin used in Google News searches.

### Supplementary Material 1. List of search terms used

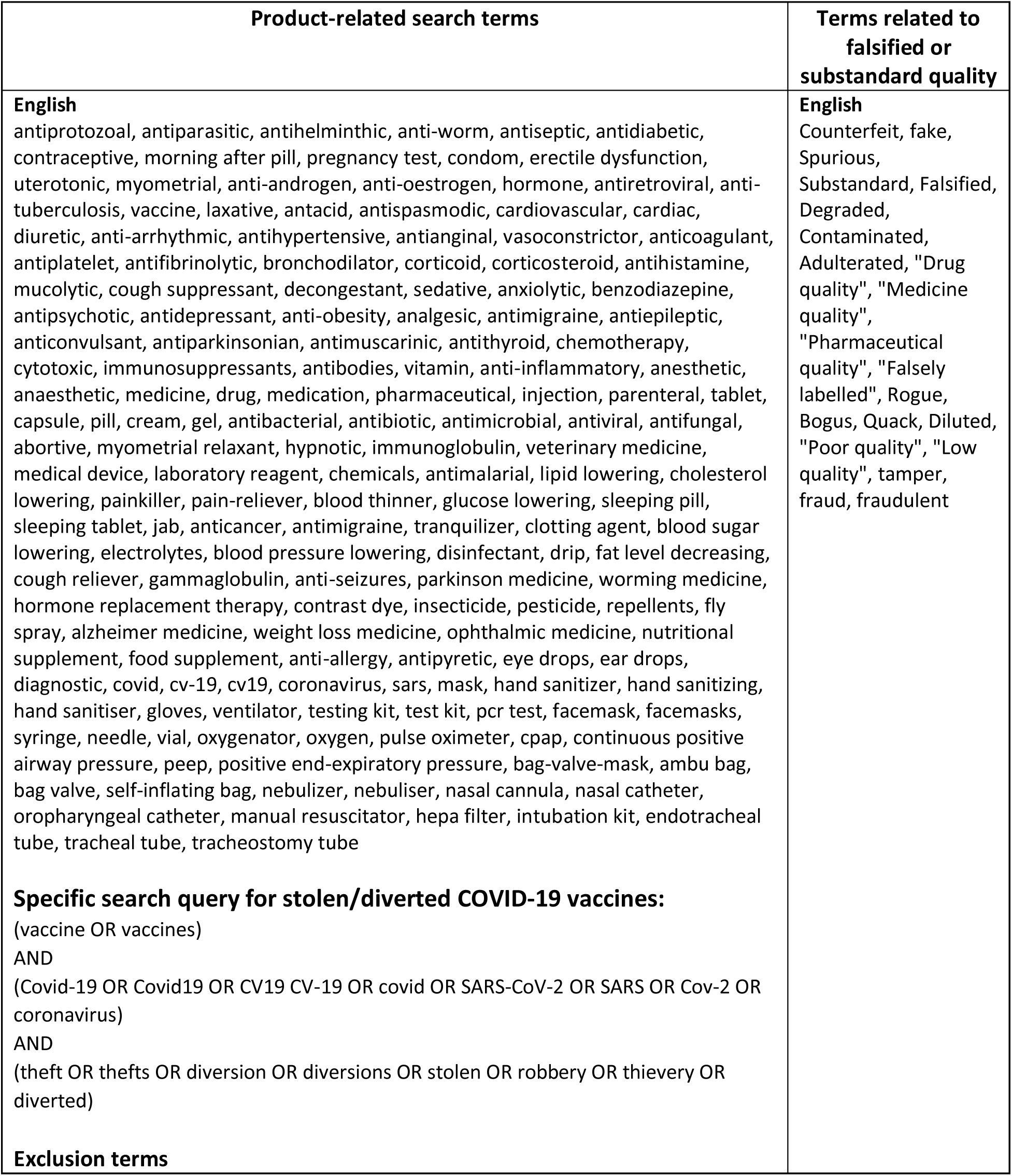
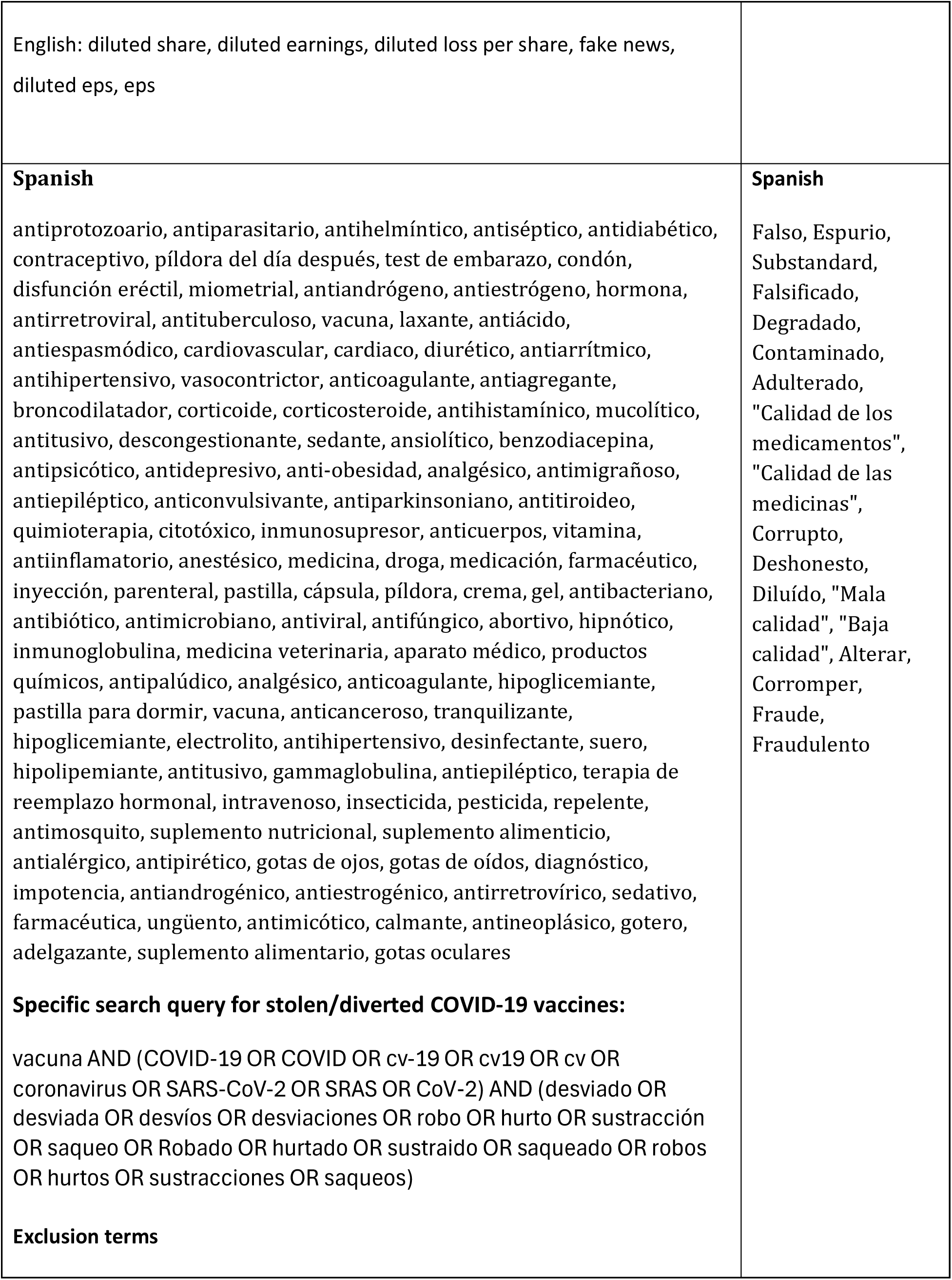
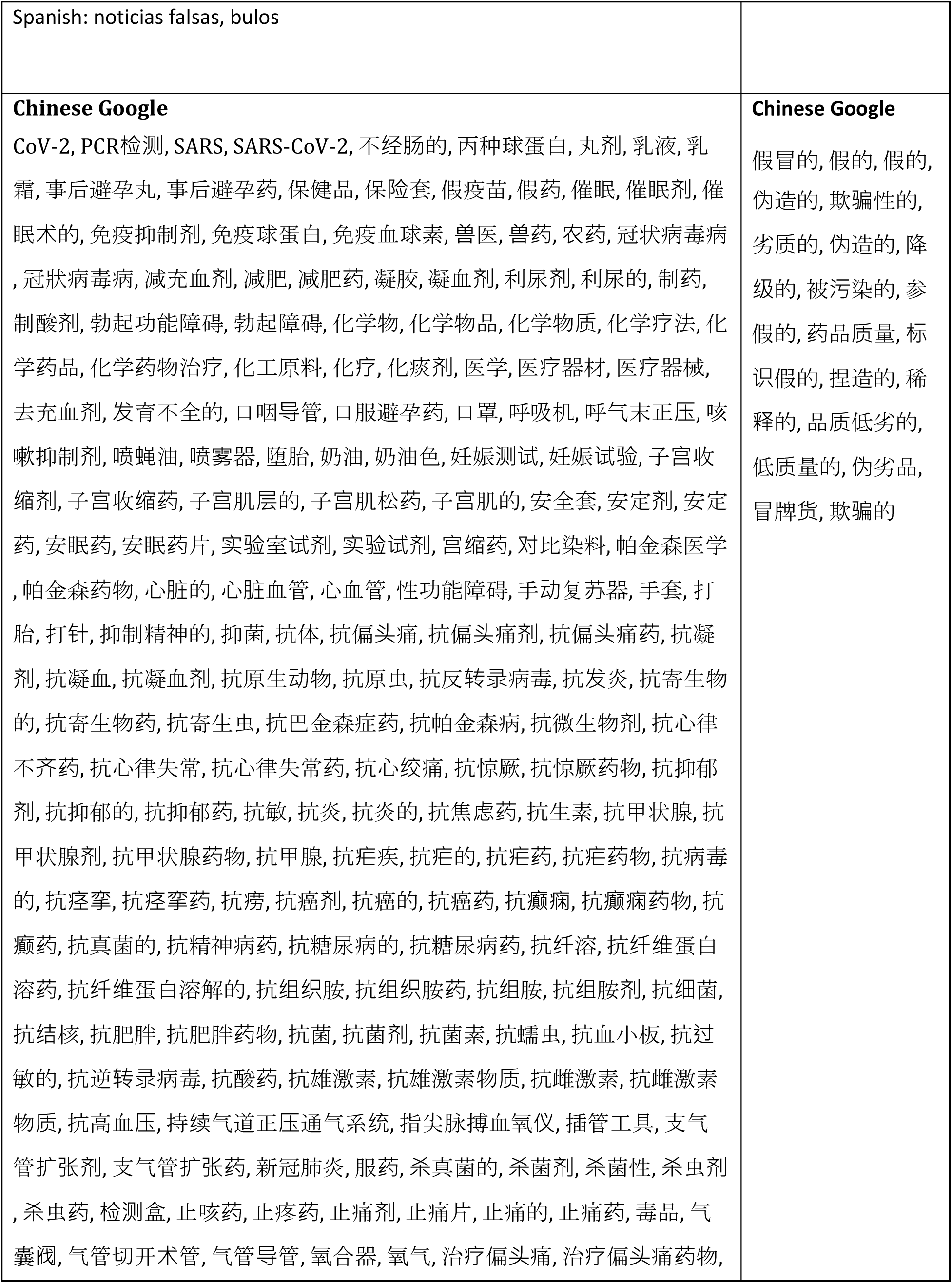
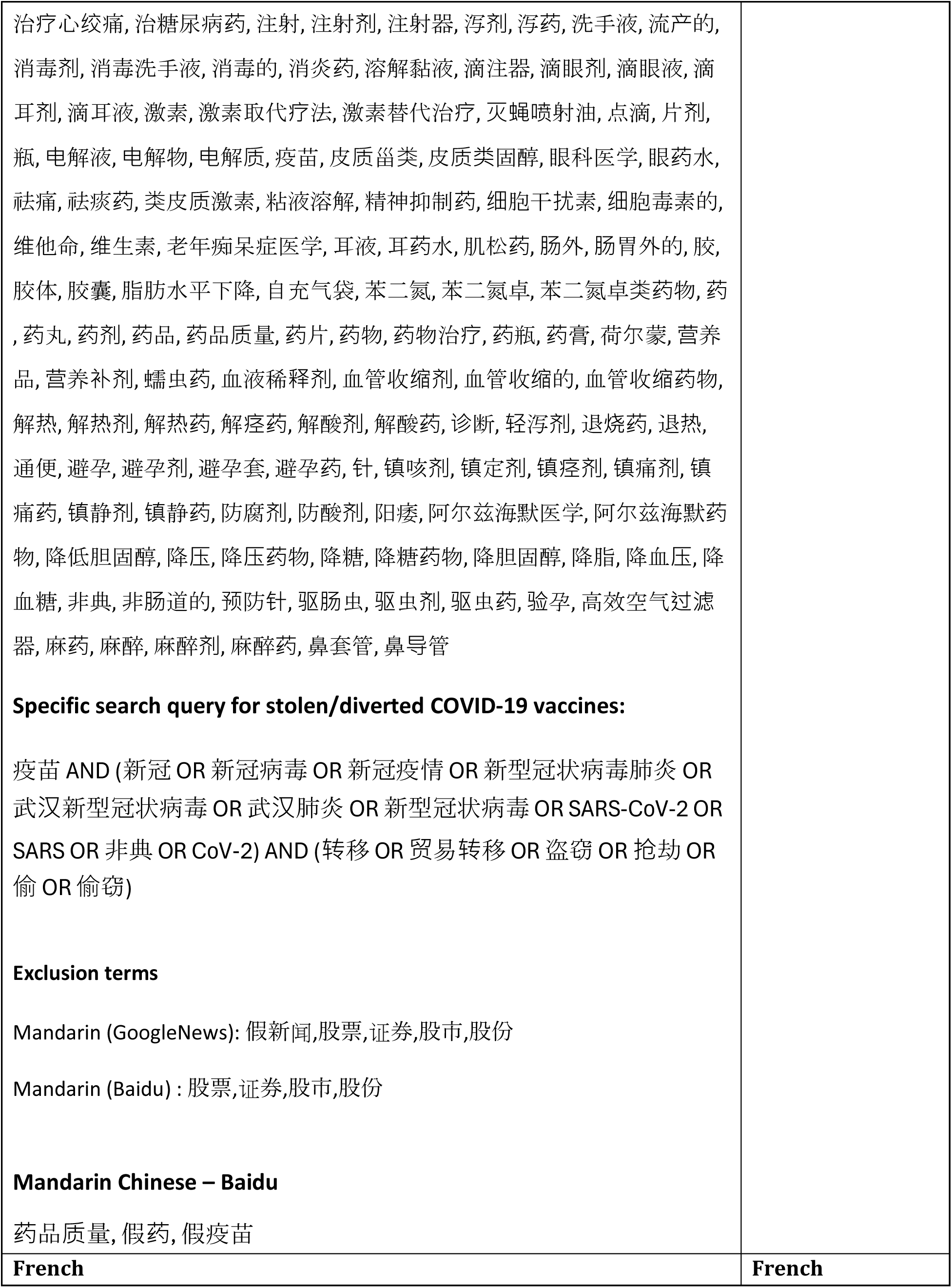
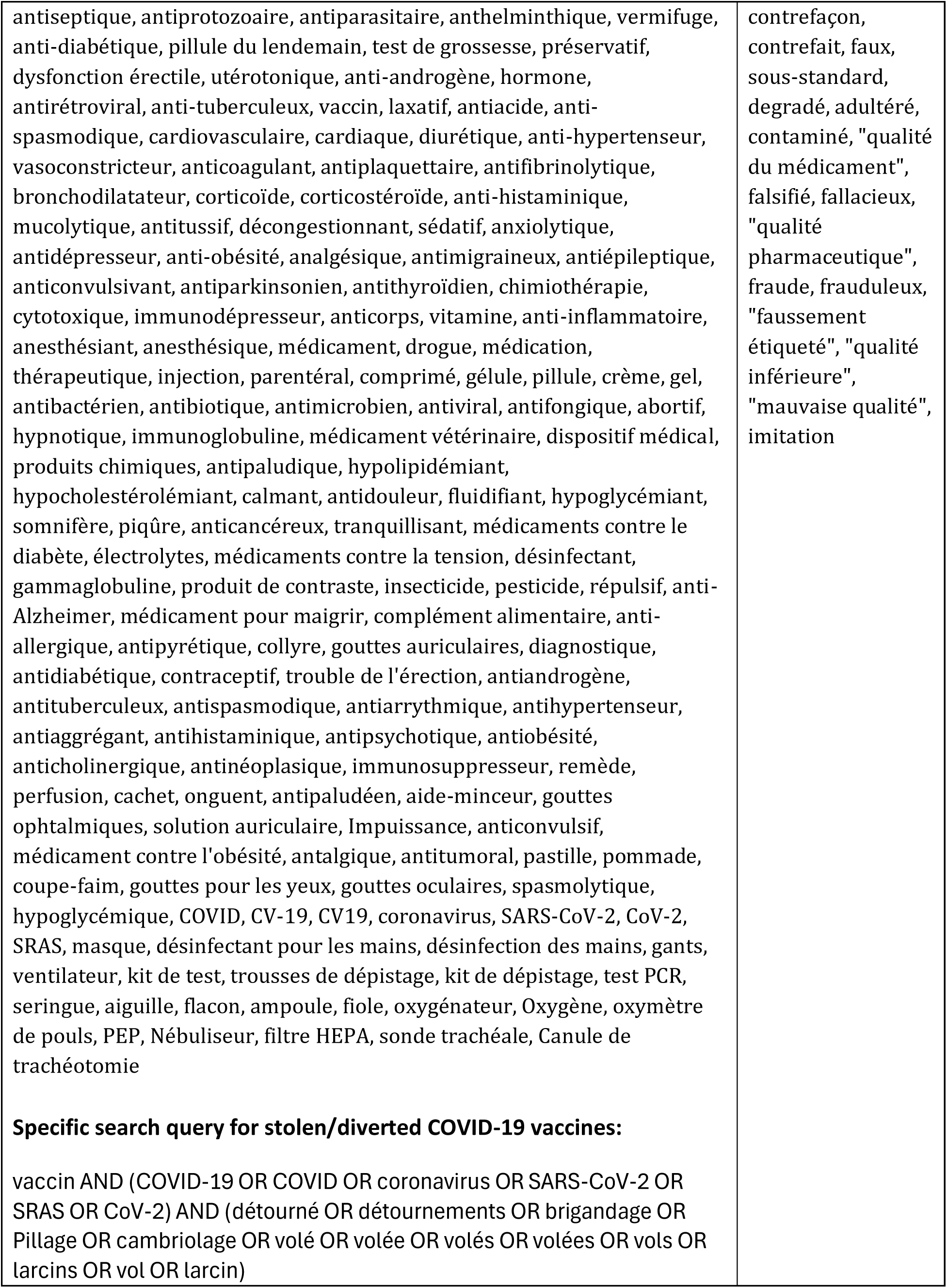
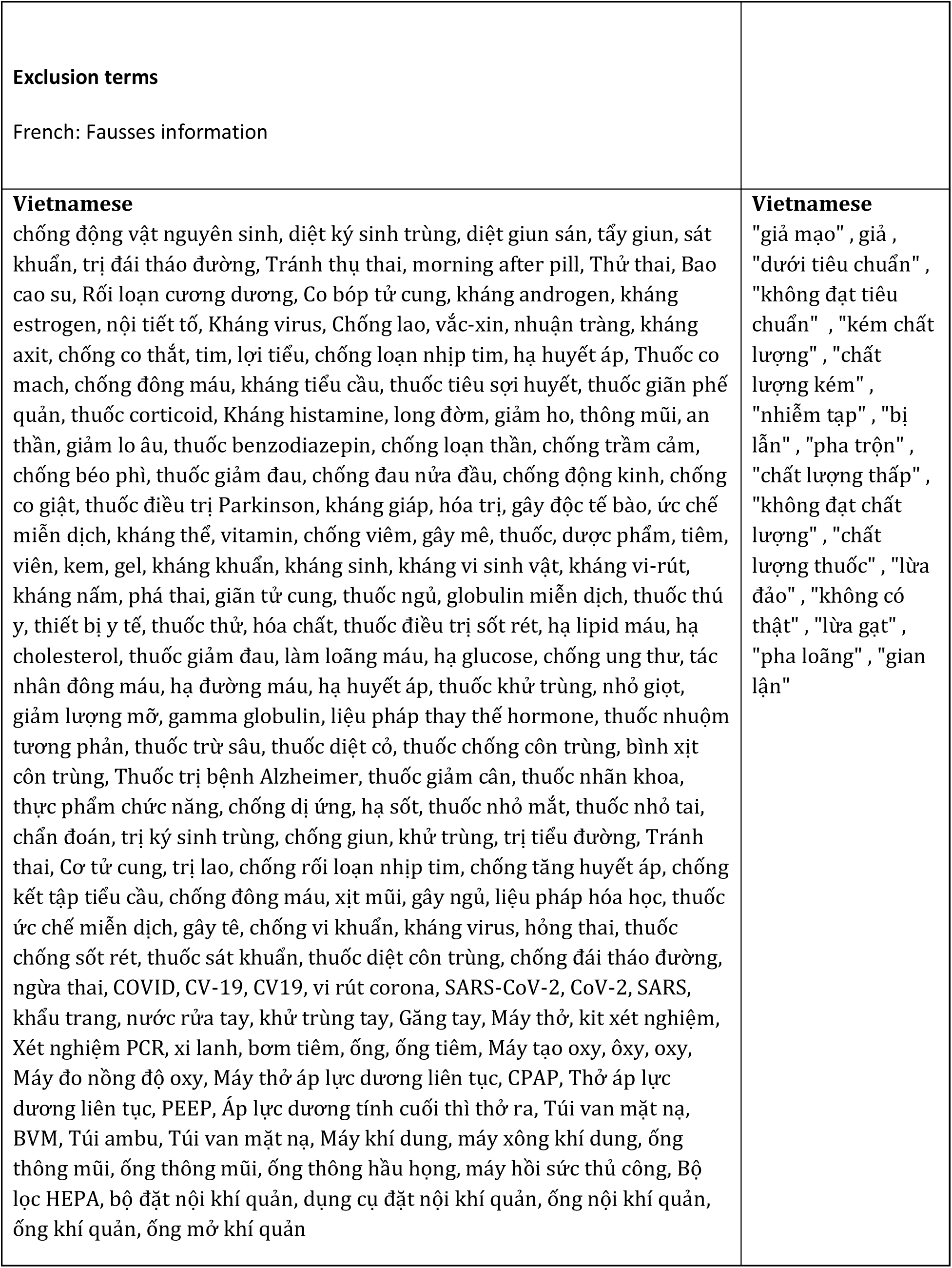
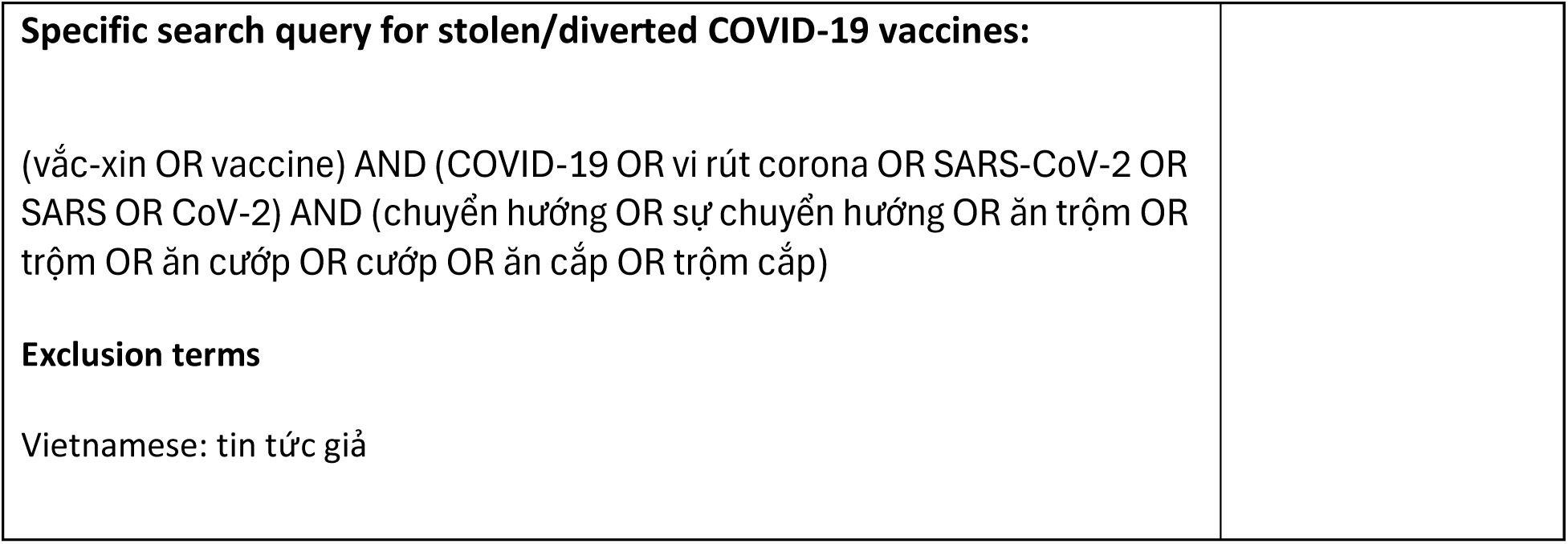

### Supplementary material 2: COVID-19 ‘Shortcut queries’ in the MQM Globe and associated key terms

Due to COVID-19 pandemic, the MQM Globe search box was adapted to enable quick access to reports of COVID medicines, Personal protective equipment (PPE), Sanitisers and disinfectant, COVID vaccines, Ventilation and Oxygenation, and COVID diagnostics.

<u>Available COVID-19 shortcuts queries on the MQM Globe web app</u>

| Shortcut name | Returns |
| --- | --- |
| COVID medicines | Medicines used to treat COVID-19 and experimental drugs in clinical trials. Includes biologics such as monoclonal antibodies. |
| Personal Protective Equipment | Gowns, gloves, facemasks, visors, etc. |
| Sanitisers and disinfectant | Wipes, disinfectant, sanitizer, etc. |
| Ventilation and oxygenation | Continuous Positive Airway Pressure, CPAP, tracheal tube, Intubation kit, etc. |
| COVID diagnostics | Thermometer, rapid diagnostic test, antigen test, lateral flow assay, etc. |
| COVID vaccines | AD26.COV2.S, Sinovac, Sputnik V, etc. |
| All COVID searches | All of the above |

<u>Key terms used for the MQM Glob web app’s COVID-19 shortcut queries</u>

1. Personal Protective Equipment ((”Personal protective equipment” OR “PPE” OR “protective glasses” OR “apron” OR “n95” OR “gowns” OR “facemask” OR “visor” OR “gloves” OR “goggles” OR “respirator” OR “KN95” OR “face shield” OR “mask”) OR ((”Medical devices for disease prevention”) AND (”COVID-19” OR “COVID” OR “SARS-CoV-2” OR “Coronavirus” OR “CV19” OR “CV-19” OR “SARS” OR “CoV-2”)))
2. COVID-19 diagnostics (”Thermometer” OR ((”coronavirus kit” OR “RDT” OR “covid test” OR “lateral flow assay” OR “test kit” OR “LFA” OR “COVID kit” OR “Medical device for screening/diagnosis/monitoring” OR “rapid diagnostic test” OR “coronavirus test” OR “antigen test” OR “COVID-19 test” OR “test cassette” OR “In-vitro-diagnostic” OR “cassette test” OR “RT-PCR” OR “IVD” OR “testing kit” OR “qPCR” OR “antibody test” OR “COVID-19 kit” OR “PCR” OR “polymerase chain reaction” OR “ELISA”) AND (”COVID-19” OR “COVID” OR “SARS-CoV-2” OR “Coronavirus” OR “CV19” OR “CV-19” OR “SARS” OR “CoV-2”)))
3. Sanitisers & disinfectants ”wipes” OR “disinfectant” OR “sanitizer” OR “sanitizing” OR “iodoform” OR “sanitiser”
4. Vaccines ((”BNT162b2” OR “BBIBP-CorV” OR “Ad26.COV2.S” OR “CoronaVac” OR “Covishield” OR “Ad5-nCoV” OR “AZD1222” OR “FBRI” OR “Sputnik V” OR “mRNA-1273” OR “EpiVacCorona” OR “Vero Cells” OR “Covaxin”) OR ((”vaccine”) AND (”Barat Biotech” OR “BioNTech” OR “Johnson & Johnson” OR “Pfizer” OR “Oxford/AstraZeneca” OR “Serum Institute of India” OR “Sinopharm” OR “Sinovac” OR “Gamaleya” OR “Moderna” OR “Pfizer/BioNTech” OR “CanSino” OR “AstraZeneca” OR “Oxford”)) OR ((”vaccine”) AND (”COVID-19” OR “COVID” OR “SARS-CoV-2” OR “Coronavirus” OR “CV19” OR “CV-19” OR “SARS” OR “CoV-2”)))
5. Ventilation & oxygenation equipment and consumables (”Continuous Positive Airway Pressure” OR “Oxygen” OR “nasal catheter “ OR “CPAP” OR “oximeter” OR “positive end-expiratory pressure” OR “PEEP” OR “positive end expiratory pressure” OR “bag-valve-mask” OR “self-inflating bag” OR “oropharyngeal catheter” OR “BMV” OR “nebulizer” OR “tracheostomy tube” OR “tracheal tube” OR “ambu bag” OR “ventilator” OR “bag valve” OR “nasal cannula” OR “manual resuscitator” OR “HEPA filter” OR “endotracheal tube” OR “air purifier” OR “intubation kit”)
6. COVID-19 medicines ”tranilast” OR “interleukin-2” OR “INC424” OR “TNKase” OR “nitazoxanide” OR “LY3832479” OR “baloxavir” OR “interleukin-7” OR “Kineret” OR “ritonavir” OR “Crizanlizumab” OR “Apixaban” OR “cyclosporin” OR “losartan” OR “ATI-450” OR “nitrogen monoxide” OR “tirofiban” OR “Ebselen” OR “corbistadine” OR “atorvastatin” OR “Eicosapentaenoic” OR “nitrite” OR “Riamilovir” OR “black cumin” OR “NK-1R” OR “Pemziviptadil” OR “colchicine” OR “Lithium” OR “Vancomycin” OR “Broncho-Vaxom” OR “ramipril” OR “Teicoplanin” OR “tofacitinib” OR “budesonide” OR “Paracetamol” OR “dipyridamole” OR “levamisole” OR “atovaquone” OR “Senicapoc” OR “covid drug” OR “enoxaparin” OR “Brequinar” OR “povidone-iodine” OR “levilimab” OR “degarelix” OR “LY3819253” OR “Sofusbovir” OR “masitinib” OR “Omega-3” OR “INM005” OR “RBT-9” OR “deferoxamine” OR “canakinumab” OR “Ramelteon” OR “chlorpromazine” OR “selinexor” OR “Piclidenoson” OR “DAS181” OR “M5049” OR “Ibudilast” OR “CM4620-IE” OR “GNS561” OR “zanubrutinib” OR “Cenicriviroc” OR “sofosbovir” OR “Trimethoprim” OR “vadadustat” OR “AVM0703” OR “Rabeprazole” OR “Moxifloxacin” OR “cobicistat” OR “BAT2020” OR “ABX464” OR “XAV-19” OR “thalidomide” OR “bamlanivimab” OR “GX-19” OR “corticosteroid” OR “Tradipitant” OR “cotrimoxazole” OR “HuMax-Inflam” OR “Apilimod” OR “DUR-928” OR “escin” OR “PF06650833” OR “octagam” OR “Antroquinonol” OR “pacritinib” OR “Imatinib” OR “ribavirin” OR “ambrisentan” OR “baricitinib” OR “imatinib” OR “CD24Fc” OR “Sulodexide” OR “AlloStim” OR “DFV890” OR “Emapalumab” OR “sitagliptin” OR “Metformin” OR “prednisone” OR “ulinastatin” OR “naltrexone” OR “abidor” OR “niclosamide” OR “BIO101” OR “GS-441524” OR “argatroban” OR “Leukine” OR “xiyanping” OR “peginterferon” OR “pembrolizumab” OR “HuMax” OR “Lambda” OR “dornase” OR “Itraconazole” OR “telemedicine” OR “Adenosine” OR “nirmatrelvir” OR “Curosurf” OR “clarithromycin” OR “bromhexine” OR “Xpovio” OR “ebastine” OR “amoxicillin/clavulanate” OR “PD-1 mAb” OR “EPA” OR “oseltamivir” OR “Betamethasone” OR “favipiravir” OR “mefloquine” OR “bismuth” OR “CM4620” OR “ifenprodil” OR “Levofloxacin” OR “REGN10987” OR “Candesartan” OR “secukinumab” OR “Trihexyphenidyl” OR “Daclatasvir” OR “pinavir” OR “tocilizumab” OR “co-amoxiclav” OR “EG-HPCP-03a” OR “hydroxychloroquine” OR “Polyoxidonium” OR “STI-5656” OR “Artesunate” OR “triazavirine” OR “Disulfiram” OR “cholecalciferol” OR “INO-4800” OR “PG1” OR “zinc” OR “oxytocin” OR “gimsilumab” OR “suramin” OR “rhG-CSF” OR “desferoxamine” OR “TD-0903” OR “OM-85” OR “Bucillamine” OR “pirfenidone” OR “Acetaminophen” OR “adamumab” OR “sulfamethoxazole” OR “BI 764198” OR “RPH-104” OR “COVID-19 drug” OR “alpha lipoic” OR “almitrine” OR “melphalan” OR “dapagliflozin” OR “NBTNM108” OR “TMJ2” OR “Icosapent” OR “Ceftriaxone” OR “isoprinosine” OR “IMU838” OR “tridecactide” OR “chloroquine” OR “CSL324” OR “Lian Hua Qing Weng” OR “Kevzara” OR “valsartan” OR “meplazumab” OR “Namilumab” OR “Prednisolone” OR “sargramostim” OR “estradiol” OR “cyclosporine” OR “Aprepitant” OR “silymarin” OR “linagliptin” OR “Noscapine” OR “Gemtuzumab” OR “methylprednisolone” OR “fluvoxamine” OR “Coroquard” OR “mavrilimumab” OR “anakinra” OR “ozanimod” OR “mepolizumab” OR “acetylsalicylic” OR “darunavir” OR “novaferon” OR “YinHu QingWen” OR “OM85” OR “camrelizumab” OR “Cosentyx” OR “estrogen” OR “dexmedetomidine” OR “LL-37” OR “Dantonic” OR “rivaroxaban” OR “adalimumab” OR “apremilast” OR “polyinosinic-polycytidylic” OR “farpiravir” OR “montelukast” OR “Ibuprofen” OR “IFX-1” OR “Iodine” OR “Molnupiravir” OR “Pioglitazone” OR “verapamil” OR “Rapamycin” OR “Brexanolone” OR “Eltrombopag” OR “ravulizumab” OR “hydrocortisone” OR “auxora” OR “tinzaparin” OR “Vascepa” OR “omalizumab” OR “Tybost” OR “Actemra” OR “dociparastat” OR “NA-831” OR “ascorbic acid” OR “MAS825” OR “C21” OR “RoActemra” OR “eculizumab” OR “Bivalirudin” OR “povidon-iodine” OR “ivermectin” OR “Pamrevlumab” OR “danoprevir” OR “Neurokinin” OR “sirolimus” OR “Fostamatinib” OR “resveratrol” OR “Icatibant” OR “bromelain” OR “dexamethasone” OR “TJ003234” OR “iloprost” OR “tacrolimus” OR “astegolimab” OR “interferon” OR “plitidepsin” OR “metenkefalin” OR “azoximer” OR “lopinavir” OR “Tazobactam” OR “carrimycin” OR “CM-4620” OR “CYT107” OR “Heparin” OR “PyronaridineArtesunate” OR “Itolizumab” OR “zilucoplan” OR “oxpentifylline” OR “AT-001” OR “Abivertinib” OR “doxycycline” OR “Nigella Sativa” OR “AZD1222” OR “leronlimab” OR “Enalapril” OR “nangibotide” OR “Piperacillin” OR “bevacizumab” OR “lactoferrin” OR “UTTR1147A” OR “Caesalpinia spinosa” OR “mometasone” OR “hydroxychloroquin” OR “Febuxostat” OR “lanadelumab” OR “Thymalfasin” OR “huaier extract” OR “Levoflozacin” OR “Pentoxifylline” OR “tozumab” OR “NP-120” OR “Alvelestat” OR “captopril” OR “merimepodib” OR “Iota-Carrageenan” OR “Lianhua Qingwen” OR “GLS-1200” OR “aescinate” OR “tranexamic” OR “Ledipasvir” OR “ISIS 721744” OR “procalcitonin” OR “SNDX-6352” OR “sirukumab” OR “Enzalutamide” OR “carriomycin” OR “amphotericin” OR “bemiparin” OR “T89” OR “Spironolactone” OR “sotrovimab” OR “fingolimod” OR “aspirin” OR “Remdesivir” OR “TJM2” OR “pyridostigmine” OR “Prolastin” OR “EC-18” OR “poractant” OR “isotretinoin” OR “telmisartan” OR “lenzilumab” OR “avdoralimab” OR “duvelisib” OR “BIO 300” OR “bicalutamide” OR “Ilaris” OR “atlizumab” OR “desferrioxamine” OR “LB1148” OR “Regkirona” OR “vitamin D3” OR “Clopidogrel” OR “CD24” OR “tetrandrine” OR “Lansoprazole” OR “Ruconest” OR “amoxicillin” OR “Trifluoperazine” OR “Ganovo” OR “nitric Oxide” OR “chlorine dioxide” OR “olokizumab” OR “lucinactant” OR “galidesivir” OR “TXA127” OR “Maraviroc” OR “conestat” OR “CA S001” OR “vazegepant” OR “REGN10933” OR “Propranolol” OR “Viagra” OR “Fisetin” OR “Ronapreve” OR “Previfenon” OR “omega 3” OR “regdanvimab” OR “thymosin” OR “Prasugrel” OR “retinoic acid” OR “Ceftaroline” OR “sevoflurane” OR “amoxicillin/clavulanic acid” OR “oestrogen” OR “leflunomide” OR “virazole” OR “PLN-74809” OR “ATYR1923” OR “Olumiant” OR “dalargin” OR “Alinia” OR “methotrexate” OR “dapansutrile” OR “artemisinin” OR “ibrutinib” OR “aescin” OR “CERC-002” OR “fludase” OR “isoflurane” OR “XPro1595” OR “LY-CoV555” OR “CAS0001” OR “immunoglobulin” OR “nafamostat” OR “Crocetinate” OR “Diphenhydramine” OR “BIO 101” OR “AZD1656” OR “PTC299” OR “amodiaquine” OR “casirivimab” OR “BGB-DXP593” OR “opaganib” OR “melatonin” OR “huaier granule” OR “HuMax-IL8” OR “Paxlovid” OR “famotidine” OR “GLS1027” OR “Trimodulin” OR “tenofovir” OR “Primaquine” OR “AMY-101” OR “covid medicine” OR “umifenovir” OR “EDP1815” OR “Lagevrio” OR “Vitamin B12” OR “Gamunex-C” OR “Bardoxolone” OR “AstroStem-V” OR “LAU-7b” OR “Vitamin E” OR “Vitamin B” OR “RTB101” OR “COVID-19 medicine” OR “curcumin” OR “fondaparinux” OR “Edoxaban” OR “L-Citrulline” OR “ciclesonide” OR “azithromycin” OR “remdesivir” OR “Diltiazem” OR “Methylene blue” OR “clazakizumab” OR “BCX4430” OR “Pyronaridine” OR “Quercetin” OR “Toremifene” OR “COVI-AMG” OR “etoposide” OR “DWJ1248” OR “defibrotide” OR “AT-527” OR “prazosin” OR “triazavirin” OR “BIO300” OR “Ensifentrine” OR “coronavirus medicine” OR “Anti-IL-8” OR “dihydroartemisinin” OR “vitamin c” OR “25-hydroxyvitamin D3” OR “coronavirus drug” OR “formoterol” OR “indomethacin” OR “Rayaldee” OR “ciclosporin” OR “naproxen” OR “fluoxetine” OR “Infliximab” OR “Tenecteplase” OR “ruxolitinib” OR “Molgramostim” OR “vitamin D” OR “simvastatin” OR “alteplase” OR “sildenafil” OR “isoquercetin” OR “GC4419” OR “ketamine” OR “Razuprotafib” OR “camostat” OR “Arbidol” OR “Montmorrillonite” OR “acalabrutinib” OR “nivolumab” OR “aviptadil” OR “PUL-042” OR “diammonium” OR “Clevudine” OR “nitrogen oxide” OR “BMS-986253” OR “siltuximab” OR “interleukin 2” OR “jakotinib” OR “nintedanib” OR “Axatilimab” OR “garadacimab” OR “Treamid” OR “ASC09” OR “emtricitabine” OR “LY-CoV016” OR “Pulmozyme” OR “Prostaglandin” OR “ciclosporine” OR “hydrogen peroxide” OR “sarilumab” OR “Losmapimod” OR “azvudine” OR “BLD-2660” OR “EIDD-2801” OR “MSTT1041A” OR “Desidustat” OR “abidole” OR “omeprazole” OR “progesterone” OR “Decitabine” OR “tocopherol” OR “berberine” OR “Xevudy” OR “APL-9” OR “colomycin” OR “XC221” OR “amiodarone” OR “lenalidomide” OR “imdevimab” OR “ixekizumab” OR “VentaProst” OR “acetylcysteine” OR “LY3127804” OR “Atazanavir” OR “TL-895” OR “dalteparin” OR “Thimerosal” OR “Xue-Bi-Jing” OR “GC376” OR “Angiotensin” OR “gs441542” OR “Risankizumab” OR “co-trimoxazole”) OR ((”Medicine” OR “Plasma” OR “Treatment” OR “Medication” OR “Monoclonal antibodies” OR “Antibody therapy” OR “Antibody cocktail”) AND (”COVID-19” OR “COVID” OR “SARS-CoV-2” OR “Coronavirus” OR “CV19” OR “CV-19” OR “SARS” OR “CoV-2”

### Supplementary material 3. Information sources for capture by direct RSS

The sources are webpages from selected MRA, specific newspapers not referenced in Google News that previously published on SF medical products, and various international organisations. RSS feeds are filtered based on a set of inclusion terms related to medicine quality and exclusion terms related to common false-positive topics.

| English |
| --- |
| US Food and Drug Administration (various newsletters) |
| Food and Drug Administration of the Philippines |
| Hindustan Times |
| Institute of Research against Counterfeit Medicines (IRACM) |
| MedicalDialogues.com, Pharmaceutical News |
| Ministry of Health, Belize |
| Modern Ghana RSS |
| National Drug Authority, Uganda |
| SecuringIndustry.com, Pharmaceuticals |
| US Customs and Border Protection News |
| Zambia Medicines Regulatory Authority |
| French |
| ANSM France |
| CAMEG, Burkina Faso |
| Centre National de Pharmacovigilance et de Matéiovigilance, Algérie |
| Direction de la Pharmacie et du Médicament, République du Sénégal |
| Direction générale de la concurrence, de la consommation et de la répression des fraudes (DGCCRF), France |
| Fondation Chirac, France |
| IRACM |
| Ministère des Solidarités et de la Santé, France |
| Ministère Economique Actualités Generales, France |
| Spanish |
| Agencia Española de Medicamentos y Productos Sanitarios |
| Ministerio de Gobierno, Ecuador |
| Ministerio de Salud Pública, Ecuador |
| Ministerio de Salud Pública, Republica Dominicana |

### Supplementary material 4. Examples of search queries illustrating the flexibility of the MQM Globe web app search function

| Search String | Meaning |
| --- | --- |
| Uganda | Searches reports for the name Uganda |
| Africa | Searches reports for the name Africa. On indexing any reports containing the name of an African country would have the name Africa added to the index. |
| Afri* | The wildcard '*' character is used to match zero or more characters. For example, Afri* will find the name Africa as well as Afrikaans. |
| Afica~ | The query parser will complete a fuzzy search on the term 'afica' and will typically find Africa |
| Africa AND Asia | Returns only those reports that contain both Africa AND Asia. N.B. the 'AND' must be in uppercase to be interpreted as a logic element. |
| snippet:impounded | Searches for the word 'impounded' only in the snippet of all the reports. |
| Vaccines NOT China | Search for all reports about vaccines that do not include details of a place in China. N.B. the 'NOT' needs to be in uppercase to be identified as a logic element. |
| benzodiazepine | The system looks for 'benzodiazepine' within the ATC table and will return any reports that contain medicines with ATC codes belonging to lower-level codes. For example, 'benzodiazepine,' found in the ATC classification as 'benzodiazepine derivatives' with ATC code "N05BA" will return reports containing 'diazepam' (N05BA01), 'chlordiazepoxide' (N05BA02), 'medazepam' (N05BA03), 'oxazepam' (N05BA04) etc. |
| "Central Africa" | Returns reports for countries in Central Africa |
| Central Africa | Returns reports for countries in Central Africa and for countries in Africa and for the word 'Central', e.g., Central Asia, etc. |
| Note: the search engine also performs stemming to remove plurals. |  |

### Supplementary material 5. Number of unique users of the Medicine Quality Monitoring Globe per year per country (top 10)

| Country | 2020 | 2021 | 2022 | 2023 | 2024 |
| --- | --- | --- | --- | --- | --- |
| United States of America | 274 | 238 | 185 | 244 | 128 |
| United Kingdom of Great Britain and Northern Ireland | 111 | 134 | 115 | 86 | 79 |
| France | 93 | 50 | 30 | 20 | 14 |
| Germany | 20 | 22 | 16 | 58 | 42 |
| India | 11 | 23 | 19 | 13 | 43 |
| Switzerland | 32 | 33 | 10 | 10 | 20 |
| Niger | 67 | 35 | 1 | 0 | 1 |
| China | 2 | 16 | 21 | 19 | 10 |
| Belgium | 18 | 13 | 9 | 11 | 7 |
| Netherlands | 20 | 13 | 12 | 7 | 5 |

